# Impact of emerging SARS-CoV-2 on total and cause-specific maternal mortality: A natural experiment in Chile during the peak of the outbreak

**DOI:** 10.1101/2024.01.17.24301426

**Authors:** Yordanis Enriquez, María Elena Critto, Ruth Weinberg, Lenin de Janon Quevedo, Aliro Galleguillos, Elard Koch

**Affiliations:** Facultad de Ciencias de la Salud, Universidad Católica Sedes Sapientiae, Lima, Peru; Facultad de Ciencias Médicas, Pontificia Universidad Católica, Buenos Aires, Argentina; Facultad de Medicina, Universidad Nacional de Buenos Aires, Buenos Aires, Argentina; Facultad de Medicina, Universidad de Chile, Santiago de Chile, Chile; MELISA Institute, Concepcion, Chile

**Author notes:** Corresponding author: Elard Koch.

**Keywords:** Maternal mortality, Infectious diseases, SARS-CoV-2, Pandemic, Maternal Health, Latin America

## Abstract

**Background:** This study estimated the effects of the SARS-CoV-2 pandemic on maternal death causes in Chile during the outbreak peak between 2020 and 2021.

**Materials and Methods:** A natural experiment was conducted using official data on maternal deaths and live births (LBs) between 1997 and 2021. Trend changes in the maternal mortality ratio (MMR) were assessed using segmented regression. The effects of the SARS-CoV-2 outbreak were evaluated using interrupted time series (ITS) and an autoregressive integrated moving average (ARIMA) model to forecast the expected rates on MMR and 95% confidence intervals (95% CI).

**Findings:** ITS analysis revealed that the SARS-CoV-2 outbreak impacted the MMR due to indirect causes, with a greater increase in indirect nonrespiratory causes than respiratory causes. The ARIMA forecast was consistent with ITS, showing that the expected MMR for indirect causes was substantially lower than the observed rates (9.65 in 2020 and 7.46/100,000 LBs in 2021). The expected MMR was 3.44 in 2020 and 1.55 in 2021. For nonrespiratory causes, the observed values of the MMR for 2020 (8.77/100.000 LBs) and 2021 (7.46/100.000 LBs) doubled the prediction 4.02 (95% CI: 0.44-7.61) and 3.83 (95% CI: -0.12-7.79). No significant effect was found on direct obstetrical deaths.

**Interpretation:** During 2020-2021, there was a rise in the MMR in Chile attributable to SARS-CoV-2. The pandemic contributed to an escalation in the MMR due to indirect causes, particularly nonrespiratory and infectious causes, suggesting that the risk of pregnant women to SARS-CoV-2 was increased from previous comorbidities.

## Introduction

At the end of January 2020, the World Health Organization (WHO) declared an outbreak of a new coronavirus (SARS-CoV-2), classifying it as a public health emergency of international importance and later a pandemic.^1^ The responses to the pandemic varied greatly among countries and were influenced by factors such as available resources, protection and prevention supplies, training and preparation of health personnel, data monitoring, health system integration, availability of critical beds and mechanical ventilators, and accessibility to medical services.^2^

In addition, despite the measures taken to prioritize access to health in the population in the face of SARS-CoV-2, a decrease in the use of medical services during the pandemic was observed in some middle- and low-income countries.^3^ The increase in mortality from SARS-CoV-2 unevenly affected vulnerable populations, such as elderly people living in nursing homes and people with preexisting diseases and comorbidities.^4^ The statistical data of each country and the scientific analysis thereof allow us to observe how the SARS-CoV-2 pandemic has had direct and indirect impacts on the health of the population worldwide, on access to services, and, in particular, on mortality.^3,5^ In this sense, the data from the mortality registers and their analysis through natural experiments allow us to establish the magnitude of the specific impact that the pandemic has had on total and cause-specific mortality.

The pandemic has notably impacted preventable deaths, including maternal deaths, thereby interrupting the progress observed in recent decades in reducing maternal mortality (MM) worldwide.^6^ In light of this, the WHO responded by issuing an epidemiological alert. This alert highlighted a higher risk of severe clinical manifestations of SARS-CoV-2 among pregnant women.^7^

Responding to the same crisis, in February 2020, the Ministry of Health of Chile decreed a Health Alert for the whole country to take measures to deal with the epidemic in its early stage.^8^ Despite having a relatively low maternal mortality ratios (MMR: 19.1/100,000 LBs in 2019) compared to other countries in the region, at the national level, the excess of MM from causes that could be attributable to SARS-CoV-2 has not yet been estimated. Such an estimate would provide not only a measure of the MM attributable to the virus but also of the effect it had on maternal mortality from specific causes.^9^ In addition, the comparison between maternal mortality data prior to the pandemic period and those attributable to the pandemic makes it possible to clarify the relevance of the impact on diverse groups of MM causes. This information is crucial for the development and execution of strategic programs and policies aimed at identifying and mitigating preventable diseases and deaths.

Given these scenario, our study proposed to assess the impact of the SARS-CoV-2 pandemic on the total and cause-specific MMR during its first two years in Chile. To achieve this, we employed a time-series design exploiting long-term annual trend information and used ARIMA models to forecast expected mortality under the hypothesis that previous mortality trends would continue in the absence of the pandemic virus-related mortality burden.

## Methods

### Study design and data sources

This was a study with an observational and ecological design using official maternal mortality data at the national level.

Time series of population data on maternal deaths from 1997 to 2021 published in database format by the Department of Health Statistics and Information (DEIS) of the Ministry of Health of Chile were prepared.^10^ The death records for 2020 and 2021 corresponded to provisional data updated by this organization on May 17, 2022. However, corrected live births came from the National Institute of Statistics of the same country, published in the Vital Statistics Yearbooks.^11^

### Study variables

The total maternal mortality ratio (MMR) was calculated as the ratio of maternal deaths (ICD-10: Codes O00 to O99, excluding codes O96 and O97) to the number of corrected live births, multiplied by 100,000. The MMR was categorized into groups of direct and indirect obstetric causes (Figure 1).

**Figure 1:**
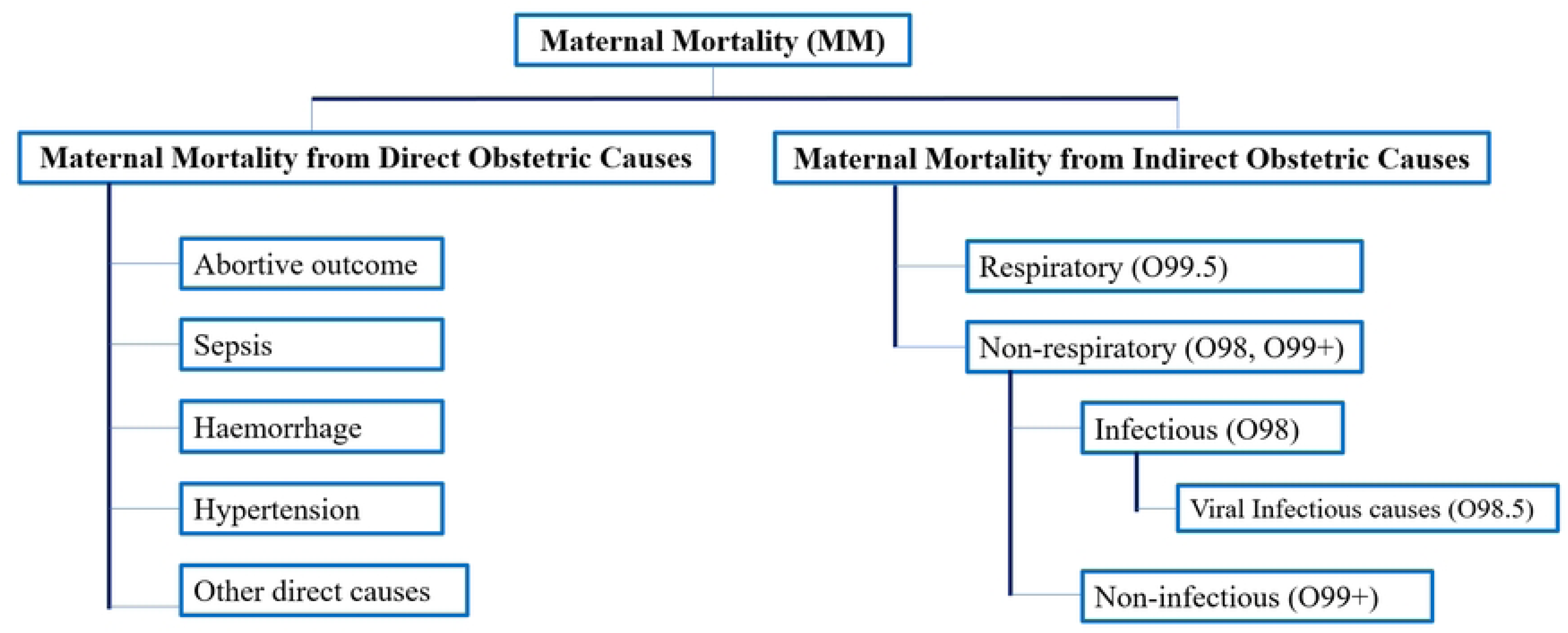
Flowchart of maternal mortality classified by cause groups

The MMR due to direct obstetric causes referred to maternal deaths resulting from obstetric complications during pregnancy, childbirth, and the postpartum period, as well as from interventions during these stages (ICD-10: Codes O00 to O99, excluding O98 and O99).^11,12^ The rate was calculated per 100,000 live births.

The MMR due to indirect obstetric causes included deaths that resulted from a preexisting disease or one that developed during pregnancy and was exacerbated by the physiological effects of pregnancy.^13^ This category of maternal deaths (ICD-10: Codes O98 and O99) per 100,000 live births was further divided into indirect respiratory and nonrespiratory obstetric causes.

The indirect respiratory obstetric MMR included maternal deaths caused by diseases of the respiratory system that were exacerbated during pregnancy, childbirth, and puerperium (ICD-10: Code O99.5). The indirect nonrespiratory obstetric MMR comprised all deaths from indirect causes, except for respiratory complications. Within this subgroup, it was possible to identify indirect infectious causes (ICD-10: O98) and indirect noninfectious causes (ICD-10: O99, except O99.5).

In Chile, following WHO suggestions, maternal deaths aggravated by SARS-CoV-2 were assigned to code O98.5 (nonrespiratory infectious indirect) accompanied by codes U07.1 and U07.2, depending on confirmation of the presence or absence of the virus.^12,13^

For the subgroups of respiratory and infectious causes of maternal mortality, due to the low frequency of events, the maternal mortality rate was amplified by 1,000,000 LBs. For the years in which no cases were reported, one case per 1,000,000 LBs was attributed.

### Statistical analysis

Analysis of MMR trends was performed with linear logarithmic regression using the Joinpoint Regression Program version 4.9.1.0 (Surveillance Research Program, National Cancer Institute) to identify the number and location of joinpoints (hereafter referred to as “joinpoints” or “junction points”) that represent the moments of trend change. In the same fashion, the estimated annual percent change (APC) of the variables for each time segment was calculated, as well as the average annual percent change (AAPC) for the entire period studied. The calculation of these estimates has already been described in detail elsewhere.^14^ The Joinpoint Regression Program does not allow analysis of dependent variables with values equal to zero for one or more years. Consequently, no results of this analysis were introduced for the MMR due to indirect respiratory causes or the MMR due to indirect infectious causes caused by the absence of maternal deaths reported for some periods.

The effect of the SARS-CoV-2 pandemic on different maternal mortality groups was first assessed using an interrupted time-series (ITS) approach in parallel trends in cause-specific mortality.

Three coefficients were estimated in this model: *β*_0_ represents a constant or starting point for the variable of interest; *β*_1_ is the change in outcome associated with an increase in the unit of time *T_t,_*which is interpreted as the preintervention trend; *β*_2_ is the level of immediate change after the *X_t_* intervention, which is specified as a dummy variable (preintervention period = 0 and postintervention period = 1); and *β*3 indicates the change in slope after the intervention with respect to the initial trend.

The ITS model considered the occurrence of a natural event such as the SARS-CoV-2 pandemic and assessed significant immediate trends and changes or sizes of effects attributable to the event of interest using the linear minimum squares method. Coefficients with standard Newey‒West errors were estimated to evaluate autocorrelation and possible heteroscedasticity.

The effect of the SARS-CoV-2 pandemic on the MMR was then estimated with an autoregressive integrated moving average (ARIMA) model.^14^ First, the stationarity of the time series was analysed using the Dickey-Fuller test. The use of the ARIMA model avoided overestimating the statistical significance of the effect of the intervention produced by autocorrelation. The effect of SARS-CoV-2 on the variables of interest was evaluated with a forecast for the years 2020 and 2021 as if this had not been verified in 2020, thus calculating an estimate of the expected MMR in the absence of the pandemic for those two years. First, for the series, a one-step-ahead training set was determined for the years 2015-2019. Then, the forecasts were compared with observed annual MMR values, identifying a difference in mortality attributable to the pandemic. For the MMR due to respiratory causes, no prediction was made for 2021, as no maternal deaths were reported in this group for this year.

Subsequently, to evaluate the accuracy of the prediction of the different models, the mean absolute error (MAE) was determined. The MAE depends on the prediction measurement scale and is therefore expressed in the same unit of measure as the analysed data, giving the size of the prediction error with an average of the difference between prediction and observation. In addition, the mean absolute percent error of prediction (MAPE) was estimated. This expresses the percentage of error attributable to the prediction and does not depend on the measurement scale of the observed data. However, this metric takes on an undefined value when zero values exist in the analysed series. The following categorization was assumed to determine the predictive ability of the models: MAPE <10 (highly accurate prediction), 10-20 (good prediction), 20-50 (reasonable prediction), and >50 (inaccurate prediction). Likewise, the percentage accuracy of the predictions was calculated by subtracting the magnitude of the error expressed in the MAPE from 100 to express the variability explained by the model. In these analyses, the software Stata version 15.1® (Stata Corp, College Station) was used, and the corresponding 95% confidence intervals (95% CIs) were estimated for the measurements.

### Ethical approval

This study used aggregated public data. The source of the information is the official maternal mortality records of Chile published by the DEIS (available at https://deis.minsal.cl/). The data was anonymised in the source. The study protocol was approved by the Ethics Committee of the Catholic University Sedes Sapientiae, Lima, Peru.

### Role of the funding source

The funder of this study had no role in the decision for publication, study design, data collection, data analysis, data interpretation, or writing of the report.

## Results

### Segmented linear regression analysis

Table 1 details the joinpoint analysis of MMR trends by cause group between 1997 and 2021. In the global MMR, a continuous downward trend was observed from 1997 to 2003, with an annual decrease of 6.09% (95% CI: -10.7 - -1.2). The first turning point in the curve was identified in 2003, while the year 2017 marked a second change in the slope. An annual growth of 2.89% was observed from 2003 to 2017 (95% CI: 1.3-4.5). As of this year, the trend decreased again annually by 5.60% (95% CI: -14.1 - -3.8) until 2021. The average annual percent change (AAPC) for the period evaluated was -0.9% (95% CI: -2.9-1.2).

**Table 1.** Segmented linear regression analyses to model trends and joinpoints for the total MMR and cause specific MMRs groups in Chile, 1997-2021.

|  | Years | APCs | p-value | 95% CI | Joinpoint | 95% CI |
| --- | --- | --- | --- | --- | --- | --- |
| MMR total |  |  |  |  |  |  |
| trend 1 | 1997-2003 | -6.09 | 0.01 | -10.7--1.2 | - | - |
| trend 2 | 2003-2017 | 2.86 | <0.001 | 1.3-4.5 | 2003 | 1999-2010 |
| trend 3 | 2017-2021 | -5.60 | 0.21 | -14.1-3.8 | 2017 | 2002-2019 |
| AAPC | 1997-2021 | -0.9 | 0.39 | -2.9-1.2 | - | - |
| MMR by direct causes |  |  |  |  |  |  |
| trend 1 | 1997-2003 | -9.24 | <0.001 | -12.8--5.5 | - | - |
| trend 2 | 2003-2010 | 1.10 | 0.57 | -2.9-5.3 | 2003 | 1999-2006 |
| trend 3 | 2010-2017 | 8.08 | 0.001 | 3.8-12.5 | 2010 | 2002-2016 |
| trend 4 | 2017-2021 | -15.29 | <0.001 | -21.5- -8.6 | 2017 | 2015-2019 |
| AAPC | 1997-2021 | -2.60 | 0.01 | -4.6--0.5 | - | - |
| MMR by indirect causes |  |  |  |  |  |  |
| trend 1 | 1997-2013 | 5.20 | 0.01 | 1.4-9.2 | - | - |
| trend 2 | 2013-2016 | -28.81 | 0.47 | -73.0-88.0 | 2013 | 2002-2015 |
| trend 3 | 2016-2021 | 25.79 | 0.04 | 1.2-56.3 | 2016 | 2014-2019 |
| AAPC | 1997-2021 | 4.0 | 0.53 | -8.1-17.4 | - | - |
| MMR by indirect respiratory causes * | - | - | - | - | - | - |
| MMR by indirect non-respiratory causes |  |  |  |  |  |  |
| trend 1 | 1997-2013 | 5.47 | 0.00 | 1.6-9.5 | - | - |
| trend 2 | 2013-2016 | -27.25 | 0.50 | -72.5-92.5 | 2013 | 2000-2015 |
| trend 3 | 2016-2021 | 24.49 | 0.04 | 0.1-54.7 | 2016 | 2012-2019 |
| AAPC | 1997-2021 | 4.20 | 0.50 | -7.8-17.8 | - | - |
| MMR by indirect infectious causes * | - | - | - | - | - | - |
| MMR by indirect non-infectious causes |  |  |  |  |  |  |
| trend 1 | 1997-2006 | 14.40 | 0.01 | 3.4-26.6 | - | - |
| trend 2 | 2006-2021 | -4.20 | 0.07 | -8.6-0.4 | 2006 | 2003-2012 |
| AAPC | 1997-2021 | 2.4 | 0.30 | -2.1-7.1 | - | - |

APC: Annual Percentage Change; AAPC: Average Annual Percentage Change; 95% CI, 95% confidence interval
\* Variable with values equal to zero for one or more years prevents analysis with Joinpoint Regression Program.

The MMR due to direct obstetric causes presented three inflection points, one in 2003, another in 2010 and the last in 2017, separating the study period into four segments. There was an annual decline of 9.24% between 1997 and 2003 and an annual growth trend of 1.10% between 2003 and 2010 (p=0,57) and 8.08% between 2010 and 2017. From this year on, the trend again decreased annually by 15.29% between 2017 and 2021. The AAPC was -2.6% (95% CI: -4.6 - - 0.5). This decrease in the AAPC was greater than that observed in the global MMR.

Two inflection points were observed for the MMR due to indirect obstetric causes in 2013 and 2016. Both points delimited the only time segment that resulted in a 28.81% reduction (95% CI: 73.0-88.0, p=0,47). An increase of 5.20% was observed (95% CI: 1.4-9.2) between 1997 and 2013, and an increase of 25.79% (95% CI: 1.2-56.3) was observed between 2016 and 2021. The AAPC for the entire period evaluated showed an increase of 4.0% (95% CI: -8.1-17.4). Among indirect obstetric causes, the nonrespiratory MMR showed two inflection points in the same years. An annual increase of 5.47% was observed between 1997 and 2013, followed by an annual decrease of 27.25% between 2013 and 2016 (p=0,50), and this trend reversed from 2016, with an annual increase of 24.49% between 2016 and 2021. The AAPC was 4.2% (95% CI: -7.8-17.8). Finally, among indirect nonrespiratory obstetric causes, noninfectious obstetric causes presented a single point of inflection in 2006, preceded by a time segment with an increase of 14.40% per year (95% CI: 3.4-26.6). The AAPC was 2.4% (95% CI: -2.1-7.1).

### Interrupted time series analysis

The effect of SARS-CoV-2 in 2020 on the diverse groups of the MMR is detailed in Table 2 and plotted in Figure 2. Thus, the interrupted time series (ITS) analysis revealed that the total MMR had a significant level change in 2020, corresponding to 3.69 (95% CI: 0.71-6.67) deaths per 100,000 LBs. However, in the period immediately after, this trend showed a significant decreasing change of 8.46 deaths per 100,000 LBs (95% CI: -8.73- -8.20) (Figure 2a).

**Figure 2:**
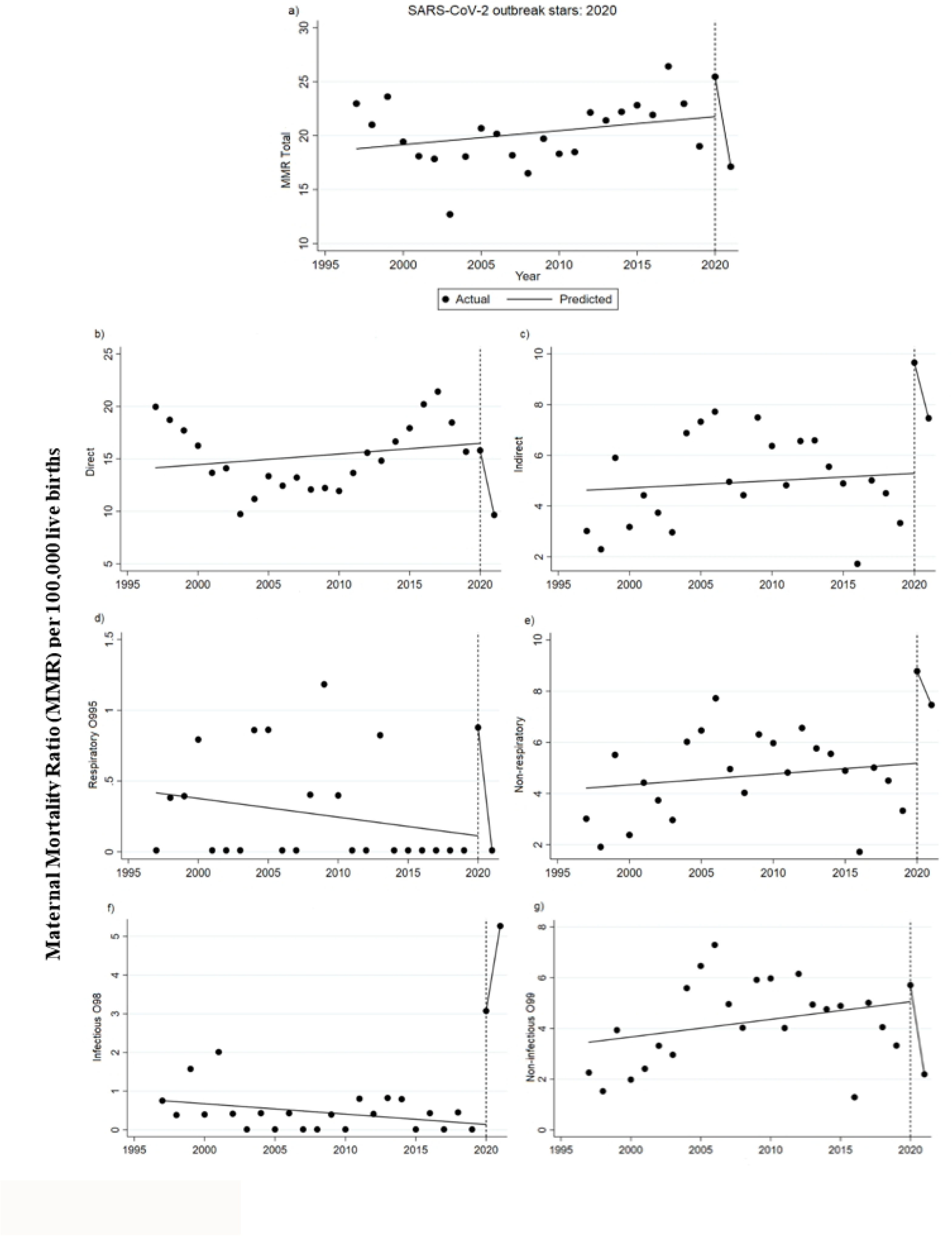
Interrupted time series analyses used to evaluate the effect of the 2020 SARS-CoV-2 pandemic on the total maternal mortality ratio (a) and maternal mortality ratios due to total direct causes (b), total indirect causes (c), respiratory causes (d), non-respiratory causes (e), infectious causes (f), non-infectious causes (g), using official records from 1997-2021. Vertical lines represent the year for the emergence of the pandemic SARS-CoV-2 influenza virus. A significant change in level due to the pandemic was identified for a, c, d, e, f, and g. However, there was no evidence of a slope change for b. An increase was observed in the following year for f, suggesting that the outbreak had an impact on mortality due to indirect causes.

**Table 2.** Effect of the SARS-CoV-2 Pandemic in 2020 on the total maternal mortality ratio (MMR) and by groups of mortality groups of causes, in Chile, 1997-2021. Interrupted time series analysis.

|  | Initial slope |  |  | Change in level |  |  | Slope change |  |  | Segment 2020-2021 |  |  |
| --- | --- | --- | --- | --- | --- | --- | --- | --- | --- | --- | --- | --- |
| | $\beta_1$ | 95% CI | p value | $\beta_2$ | 95% CI | p-value | $\beta_3$ | 95% CI | p value | $\beta_4$ | 95% CI | p value |
| MMR total | 0.13 | -0.13-0.39 | 0.32 | 3.69 | 0.71-6.67 | 0.01 | -8.46 | -8.73- -8.20 | <0.000 | -8.33 | - | - |
| MMR by groups of causes |  |  |  |  |  |  |  |  |  |  |  |  |
| Direct causes | 0.10 | -0.25- 0.46 | 0.56 | -0.67 | -5.04-3.70 | 0.75 | -6.24 | -6.60- -5.88 | <0.000 | -6.14 | -6.14 - -6.15 | <0.000 |
| Indirect causes | 0.02 | -0.10-0.16 | 0.66 | 4.36 | 2.48-6.24 | <0.000 | -2.22 | -2.35- -2.08 | <0.000 | -2.19 | - | - |
| Indirect respiratory causes | -0.01 | -0.03- -0.00 | 0.15 | 0.76 | 0.51-1.01 | <0.000 | -0.85 | -0.87- -0.83 | <0.000 | -0.86 | -0.87- -0.86 | <0.000 |
| Indirect non-respiratory causes | 0.04 | -0.07-0.16 | 0.24 | 3.59 | 1.91-5.27 | <0.000 | -1.35 | -1.47- -1.23 | <0.000 | -1.31 | - | - |
| Indirect infectious causes | -0.02 | -0.05-0.00 | 0.06 | 2.93 | 2.63-3.23 | <0.000 | 2.22 | 2.19-2.25 | <0.000 | 2.19 | 2.19-2.20 | <0.000 |
| Indirect non-infectious causes | 0.69 | -0.05-0.19 | 0.27 | 0.65 | -1.00-2.31 | 0.42 | -3.58 | -3.70-3.45 | <0.000 | -3.51 | 3.51-3.52 | <0.000 |

MMR: Maternal Mortality Ratio per 100,00 live birth; $\beta_1$ : pending before the intervention; $\beta_2$ : level change in the period immediately after the start of the intervention; $\beta_3$ : interaction term between the time from the start of the study and the variable that represents the pre- and post-intervention periods, indicating an effect over time; $\beta_4$ : calculated trend of the model through the linear combination of coefficients ( $\beta_1 + \beta_3$ ).

Regarding the direct-cause MMRs, the only significant segment was in the period after 2020, with -6.24 deaths per 100,000 LBs (95% CI: -6.60- -5.88) (Figure 2b). In contrast, the MMR due to indirect obstetric causes reported a significant level change of 4.36 deaths per 100,000 LBs (95% CI: 2.48-6.24) in 2020 (Figure 2c). Then, in the period immediately after, the trend decreased by 2.22 deaths per 100,000 LBs (95% CI: -2.35- -2.08).

Within the indirect MMR, the group of indirect nonrespiratory causes showed a greater level change in 2020 (Figure 2e) compared to the group of indirect respiratory causes (Figure 2d) (3.59 per 100,000 LBs vs. 0.76 per 100,000, respectively). However, in the period after 2020, both groups of indirect maternal deaths showed a significant decrease in the trend, being of greater magnitude in the group of nonrespiratory causes (1.35 per 100,000 LBs vs. 0.85 per 100,000, respectively).

On the other hand, the MMR due to indirect infectious causes reported statistically significant changes in the trend, with a significant change in level in 2020 of 2.93 deaths per 100,000 LBs (95% CI: 2.63-3.23), followed by an annual increase of 2.22 deaths per 100,000 LBs (95% CI: 2.19-2.25), in the period immediately after the onset of the pandemic (Figure 2f). In a similar vein, the MMR due to indirect noninfectious causes showed a significant decreasing change only in the period after 2020, with 3.58 deaths per 100,000 LBs (95% CI: -3.70-3.45) (Figure 2g).

### Forecast of the MMR with ARIMA models

The graphical inspection of the time series indicated the presence of trends in all groups of MMRs. In this way, the seasonality and subsequent differentiation of the time series of the mortality groups were verified. Supplementary Table S1 reports the parameters considered for the assessment of the models selected for the forecasts. For this purpose, the highest logarithm of likelihood, the lowest Akaike information criterion (AIC), and the lowest Bayesian information criterion (BIC) were considered for each model.

Based on the univariate ARIMA analysis, adjusted for the preinstallation data of the SARS-CoV- 2 pandemic, a forecast of expected cases was developed for 2020 and 2021. The observed and predicted values are detailed in Table 3 and plotted in Figure 3. The prediction reflected the MMR that might have been reported if the pandemic had not occurred. In 2020 and 2021, MMRs of 25.46 and 17.12 per 100,000 LBs were reported, respectively. The prediction for the first year showed 19.18 maternal deaths per 100,000 live births (95% CI: 13.41-24.95) and 17.77 (95% CI: 8.85-26.70) for 2021 (Figure 3a). Consequently, for the first year (2020), the maternal mortality difference attributable to the pandemic was 6.28 deaths per 100,000 live births. Among the possible candidates, the best model identified for the total MMR had the components (4,2,0). The accuracy of this prediction was 89.75%, with an MAE of 1.98 maternal deaths per 100,000 LBs. However, for 2021, the value of the prediction was slightly higher than that observed (see Table 3).

**Figure 3:**
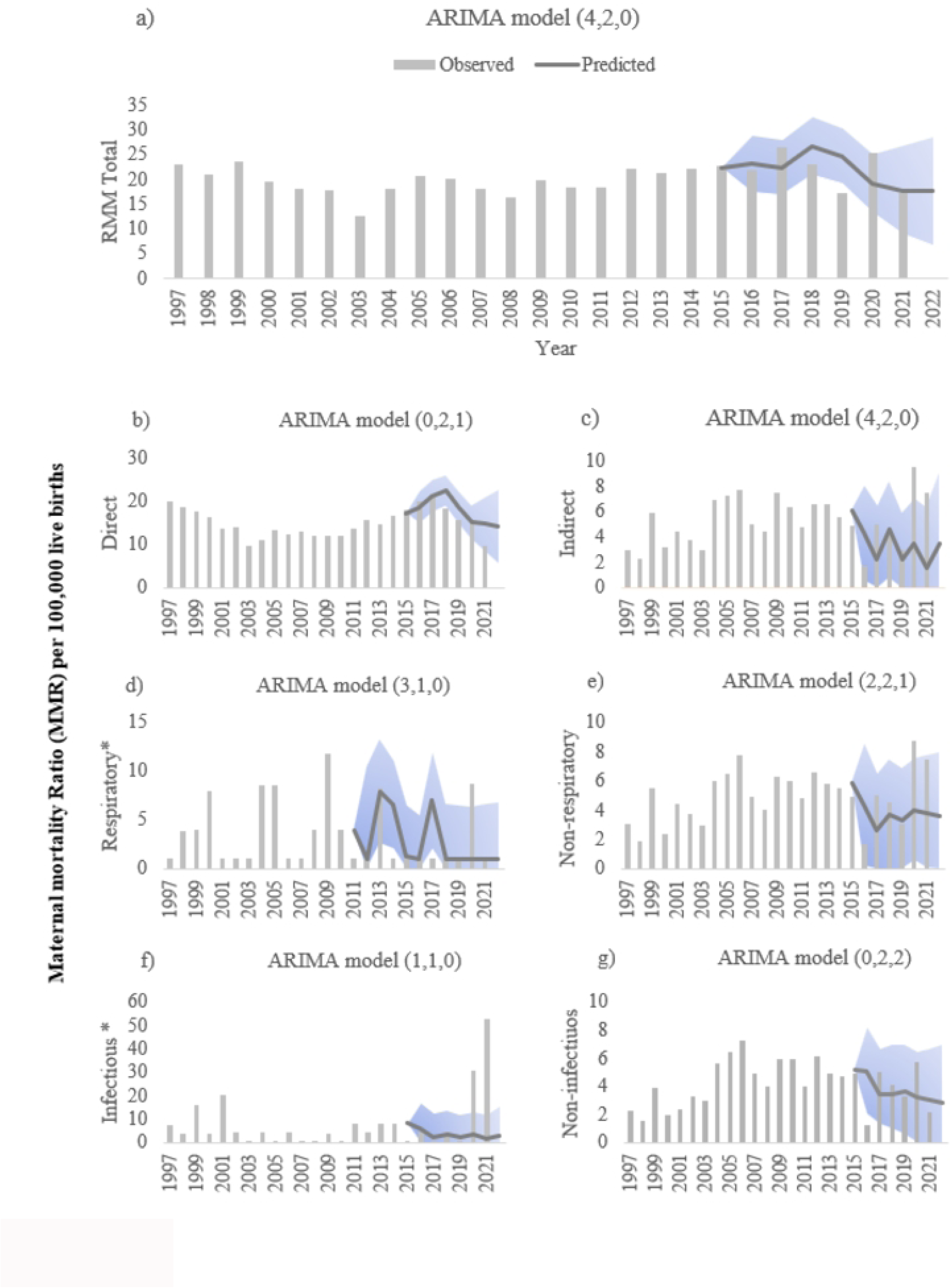
Expected maternal mortality ratio vs observed during the SARS-Cov-2 pandemic in Chile for (a) total maternal mortality ratio, (b) maternal mortality ratios due to total direct causes, (c) total indirect causes, (d) respiratory causes, (e) non-respiratory causes, (f) infectious causes, (g) non-infectious causes. * Maternal mortality ratio (MMR) per 1 000,000 live births. The line represents predicted values of the ARIMA models of MMR in Chile for total and specifics causes of maternal mortality ratio. Bars represents historical data. Light grey represents confidence intervals (CI 95%) for predicted MMR based on historical trends. For indirect causes all panels show an increase in MMR compared to expected rates based on historical data, however MMR due to direct cause decrease as seen in Panel b.

**Table 3.** Impact of the SARS-CoV-2 Pandemic on the total maternal mortality ratio (MMR) and by groups of causes of mortality, in Chile (1997-2021). Observed values versus prediction without Pandemic. Forecast analysis with ARIMA models.

|  | Year | Observed | Predictio<br>n | 95% CI | Delta | (p, d, q)<br>a | MAPE<sup>b</sup> |
| --- | --- | --- | --- | --- | --- | --- | --- |
| MMR Total | 2020 | 25.46 | 19.18 | 13.41-24.95 | 6.28 | (4, 2, 0) | 10.25 |
|  | 2021 | 17.12 | 17.77 | 8.85-26.70 | -0.65 |  |  |
|  | 2022 | - | 17.57 | 6.69-28.45 |  |  |  |
| RMM by groups of causes |  |  |  |  |  |  |  |
| Direct causes | 2020 | 15.80 | 15.22 | 11.28-19.15 | 0.58 | (0, 2, 1) | 10.33 |
|  | 2021 | 9.65 | 14.76 | 8.54-20.98 | -5.11 |  |  |
| Indirect causes | 2020 | 9.65 | 3.44 | -0.13-7.01 | 6.21 | (4, 2, 0) | 27.27 |
|  | 2021 | 7.46 | 1.55 | -3.24-6.34 | 5.91 |  |  |
| Indirect respiratory causes* | 2020 | 8.80 | 1.00 | -4.30-6.30 | 7.80 | (3, 1, 0) | 109.33 |
|  | 2021 | 0.00 |  |  |  |  |  |
| Indirect no respiratory causes | 2020 | 8.77 | 4.02 | 0.44-7.61 | 4.75 | (2, 2, 1) | 34.63 |
|  | 2021 | 7.46 | 3.83 | -0.12-7.79 | 3.63 |  |  |
| Indirect infectious causes* | 2020 | 30.70 | 3.45 | -6.15-13.06 | 27.25 | (1, 1, 0) | 154.51 |
|  | 2021 | 52.61 | 1.73 | -8.29-11.76 | 50.88 |  |  |
| Indirect non-infectious causes | 2020 | 5.70 | 3.23 | -0.02-6.5 | 2.47 | (0, 2, 2) | 35.16 |
|  | 2021 | 2.19 | 2.79 | -0.64-6.6 | -0.82 |  |  |

95% CI, 95% confidence interval; <sup>a</sup> Model components, where p represents the largest number of lags of the autoregressive parameter, d is the degree of differentiation of the series, and q is the largest number of components of the moving average; <sup>b</sup> MAPE: Mean Absolute Percent Error of prediction.
\*The estimate assumes an indefinite value when there are values equal to zero in the analyzed series.

On the other hand, for 2020 and 2021, the direct-cause MMRs were 15.22 and 14.76 per 100,000 LBs, respectively. In 2020, the ARIMA model prediction of maternal deaths per 100,000 live births for this group was 15.22 (95% CI: 11.28-19.15) and 14.76 (95% CI: 8.54-20.98) for 2021 (Figure 3b). Thus, the difference in maternal mortality attributable to the pandemic for 2020 was 0.58 deaths per 100,000 live births. However, for 2021, the observed MMR value for this group of causes (9.65 per 100,000 LBs) was lower than the prediction (see Table 3). The prediction accuracy for direct causes was 89.67%, and the model had the components (0,2,1), with an MAE of 1.43 maternal deaths per 100,000 LBs.

The best model for the MMR due to indirect causes was (4,2,0). The MMRs in this group were 9.65 and 7.46 per 100,000 LBs for 2020 and 2021, respectively. In contrast, the ARIMA model predictions for the same period were 3.44 (95% CI: -0.13-7.01) and 1.55 (95% CI: -3.24-6.34) (Figure 3c). In the comparison between groups of direct and indirect causes of death, this group had the greatest difference attributable to the pandemic, with an MMR of 6.21 per 100,000 LBs in 2020 and 5.91 in 2021, with an accuracy of 72.73% and an MAE of 1.16 maternal deaths per 100,000 LBs.

Regarding the group of indirect nonrespiratory causes, the observed values of maternal mortality in 2020 (MMR 8.77/100,000 LBs) and in 2021 (MMR 7.46/100,000 LBs) almost doubled the prediction. In this regard, the forecasts in the absence of a pandemic were 4.02 (95% CI: 0.44- 7.61) and 3.83 deaths per 100,000 LBs, respectively (Figure 3e). The difference in maternal mortality attributable to the SARS-CoV-2 pandemic would be an MMR of 4.75 (2020) and 3.63 maternal deaths per 100,000 LBs in 2021. The best model for this group of MMRs was (2,2,1), while the ARIMA model prediction for the time series had an accuracy of 65.37%.

Finally, in the group of indirect infectious causes of mortality, the MMR was 30.70/1,000,000 LBs in 2020, with an increase in the following year reaching 52.61/1,000,000 LBs. The prediction for 2020 was 3.45 (95% CI: -6.15-13.06) maternal deaths/1 000 000 LBs, and for 2021, it was 1.73 (95% CI: -8.29-11.76) maternal deaths//1 000 000 LBs (Figure 3f). Thus, the increase in mortality attributable to the pandemic would be 27.25 maternal deaths/1 000 000 LBs in 2020 and 50.88 maternal deaths/1,000,000 LBs in 2021. In the prediction, the best model for this series had the parameters (1,1,0) and an MAE of 3.22 maternal deaths per 1,000,000 LBs. The information for the other groups evaluated is shown in Table 3.

## Discussion

In order to identify the excess maternal mortality associated with the SARS-CoV-2 pandemic, this population-based study used ARIMA predictive models to estimate expected death rates in the absence of the pandemic event. Using this analytic approach, an increase in the observed total MMR for 2020-2021 relative to the expected values was confirmed. The finding was consistent with the results of an interrupted time series analysis for the secular trend. The latter showed that SARS-CoV-2 pandemic had an increasing effect on maternal mortality in Chile. There was a specific increase in the MMR due to indirect causes, specifically due to non-respiratory, infectious, and some respiratory causes. In contrast, there was no increase in the MMR due to direct obstetric causes during 2020-2021 showing a steadily downward trend over the last years.

The findings in Chile confirm preliminary surveillance reports during the outbreak. A Mexican study reported an increase in the MMR of 11.3/100,000 LBs in 2020 (42.4/100,000 LBs in 2020 compared to an MMR of 31.1/100,000 LBs in 2019).^15^ Other studies reported an increase in maternal mortality associated with the pandemic in Brazil ^16,17^ and Colombia.^6^ In this regard, some factors that would explain this increase could be difficulties in accessing maternal health care and controls, as well as interruptions in specialized services, mobility restrictions, and other diseases that were not treated in time, particularly for high-risk pregnancies.^18–21^ Likewise, the role of the fewer resources allocated to health, together with economic inequalities, may have increased the risk of maternal death from SARS-CoV-2.^17^ It is expected that difficulties in accessing health care among pregnant women would have an impact on their health and, therefore, on mortality from direct and indirect causes.^22^ However, from 2020-2021, no excess maternal mortality from direct obstetric causes associated with the pandemic was observed in Chile. This matches the results of a natural experiment conducted in Argentina with maternal mortality data (1980-2017).^14^ The study found no effect of the H1N1 pandemic on the MMR due to direct causes such as hypertension, haemorrhage, abortion outcomes, or other direct obstetric causes.

A decrease in the MMR due to direct causes was observed in 2021, which could explain the negative delta of the total MMR in the ARIMA analysis for the same year. Coincidentally, ITS analysis showed no effect in this group of direct causes. The negative difference between prediction and observation could be explained by a possible reactivation of the health system and routine obstetric controls associated with a decrease in the MMR.^8^ In this way, from 2020-2021, both the public and private health systems of Chile implemented coordinated and monitored daily health strategies focused on guaranteeing quality care for pregnant women in situations of greater vulnerability, ensuring prenatal check-ups and postnatal care, and access to emergency obstetric services. Another explanatory hypothesis, however, would be the possible underreporting of maternal deaths due to the provisional nature of the data for 2021.

On the other hand, both analyses identified groups of indirect causes, particularly non-respiratory causes in Chile for 2020 and 2021. This was consistent with other studies.^21,23–25^ Indirect causes are associated to non-transmissible comorbidities, such as diabetes, asthma, and obesity and it is well known that COVID-19 impacted predominantly on groups with this preexisting conditions. Other studies reported indirect respiratory causes as a relevant group in contributing to maternal mortality. Acute respiratory distress syndrome and pneumonia associated with SARS-CoV-2 have been indicated as leading causes of death among pregnant women.^21^ The Mexican study reported 32% increase in the group of MMRs due to indirect respiratory causes compared to the previous period (2011-2019).^15^ In regard to previous studies on the effect of pandemic events, such as the case of Argentina for emergent H1N1 in 2009, a specific increase in maternal mortality due to respiratory causes was observed.^14^ In contrast, our study identified only a slight increase in indirect respiratory causes. One hypothesis that could explain these differences would be the criterion for coding the causes of maternal deaths.

In Chile, following WHO suggestions, maternal deaths aggravated by SARS-CoV-2 are assigned to code O98.5 (non-respiratory infectious indirect) accompanied by code U07.1 or U07.2, depending on confirmation of the presence or absence of the virus.^12^ In this sense, in 2021, the group of causes of maternal deaths that had the greatest increase compared to 2020 was the group that included the deaths assigned to these codes. This increase in maternal mortality from SARS-CoV-2 in Chile in 2021 compared to 2020 coincides with the findings of a study carried out in Brazil, where it was observed that, in 2021, maternal mortality increased with the arrival of the Gamma variant.^26^

Physiological changes during pregnancy may favour complications in the presence of SARS- CoV-2.^27^ The change from cellular to humoral immunity during pregnancy is related to increased susceptibility to viral infections in pregnant women.^28^ At present, however, the information available on the increased susceptibility to SARS-CoV-2 during pregnancy is contradictory. In fact, it has been proposed that pregnant women, in relation to the general population, may have a lower morbidity and mortality.^21^ In this sense, it has been suggested that human chorionic gonadotropin and progesterone lower the proinflammatory activity of TH-1, decreasing tumour necrosis factor.^21^ This modulating effect is hypothesized to protect against the cytokine storm and, therefore, mortality associated with SARS-CoV-2 during pregnancy.^29^ However, the burden of pre-existing chronic diseases could nullify this hypothetical protective factor during pregnancy. Thus, according to the findings of a study in Brazil, pregnancy and postpartum may be important risk factors associated with severe COVID-19.^26,30^

A fresh perspective in this discussion, considering that the MMR from direct obstetric causes has remained unchanged, suggests that the pathological pathways of this emerging coronavirus are largely unrelated to pregnancy itself. Our findings indicate that SARS-CoV-2 primarily affects women with pre-existing conditions and comorbidities such as diabetes and obesity. Therefore, it is likely that this new coronavirus is acting on systems and organs already compromised by pre-existing chronic conditions, rather than the physiological changes induced by the temporary state of pregnancy.

The strengths of this natural experiment include the use of two robust statistical analysis techniques, ITS and prediction with ARIMA models. The first allowed us to determine the causal effect of a specific factor by controlling for the temporal trend before and after the event of interest. Second, we aimed to forecast and estimate the MMR in the absence of the SARS-CoV-2 pandemic with good accuracy levels. Similarly, population-based studies reduce the different sampling biases typical of observational studies, under the condition that the entire population of pregnant women exposed or not exposed to the SARS-CoV-2 pandemic is included, thus covering all deaths potentially attributable to it. In addition, according to our knowledge, this is the first Chilean study to consider at the national level the impact of the pandemic on maternal mortality and to distinguish groups of causes of mortality with the ICD-10 classification. This provides relevant epidemiological information since population-based estimates of maternal mortality in Latin America during the pandemic period are insufficient.^31^

On the other hand, among the limitations is that the data on the MMR in Chile, currently published by the Ministry of Health for the years 2020-2021, are provisional and susceptible to further adjustments. In addition, the level of certainty of prediction values may have been affected by the absence of data for some years. However, two error measures, the mean absolute error and mean absolute percentage error, were estimated to give a measure of a good level of accuracy. Likewise, multiple factors were not controlled for, including the number of pregnant women who received SARS-CoV-2 vaccines and the number of beds and respirators available during this period, so the results should be interpreted with caution.

In conclusion, the Chilean maternal mortality registry was revealed as a useful tool to evaluate the specific effects of SARS-CoV-2 on the health of pregnant women. In Chile, from 2020-2021, there was an increase in total maternal mortality associated with SARS-CoV-2. In particular, the pandemic contributed to an increase in the MMR due to indirect causes, specifically, non-respiratory and infectious causes. However, there was no effect on the MMR due to direct obstetric causes. This suggests that the increase in maternal mortality occurred mainly among women with previous comorbidities. It is imperative, then, to prevent damage to maternal and child health during epidemic periods and that health strategies be implemented that focus on ensuring continuity of quality care, ensuring recommended prenatal and postnatal check-ups and access to specialized obstetric services for high-risk pregnancies.

The results also point to the need to strengthen the surveillance system for vulnerable populations, such as pregnant women, and thus provide an integrated timely response from health systems to mitigate specific adverse effects on maternal mortality both in Chile and in other Latin American countries. Identifying and analyzing the specific causes of maternal death associated with COVID-19 could facilitate the development of strategies to prevent potential pregnancy-related deaths during future pandemics. Finally, the quality and timeliness of official cause-specific maternal mortality data are essential for timely evidence-based decision-making, especially during health crises. In this sense, Chilean official records are sensitive to natural events such as the current pandemic, and their analysis is strategic to prevent preventable maternal mortality and improve women’s health.

## Conflict of interest

The authors have no conflict of interest with the results or conclusions of this study.

## Data sharing statement

The data underlying this article were accessed from Departamento de Estadísticas de Información de Salud del Gobierno de Chile, https://deis.minsal.cl/. The derived data generated in this research will be shared on reasonable request to the corresponding author.

## Funding

This study was supported by research grants EPI-092018-01 and ONE-052021-01, both granted by FISAR http://www.fisarchile.org/.

## Supporting information - Captions

Table S1 Characteristics of the ARIMA models, selection criteria and accuracy indicators of the prediction of the maternal mortality ratio (MMR) groups of mortality, in Chile (1997-2021)

## Data Availability

The data underlying this article is available from Departamento de Estadísticas de Información de Salud del Gobierno de Chile, https://deis.minsal.cl/. The derived data generated in this research will be shared on reasonable request to the corresponding author.

## Notes

**Funding** This study was supported by FISAR.

### Competing Interest Statement

The authors have declared no competing interest.

### Author Declarations

This was a secondary data analysis of publicly available information. The source of the information is the official anonymous maternal mortality records of Chile published by the DEIS (available at https://deis.minsal.cl/). The study protocol was approved by the Ethics Committee of the Catholic University Sedes Sapientiae, Lima, Peru.

